# Anti-SARS-CoV-2 IgG responses are powerful predicting signatures for the outcome of COVID-19 patients

**DOI:** 10.1101/2020.11.10.20228890

**Authors:** Qing Lei, Cai-zheng Yu, Yang Li, Hong-yan Hou, Zhao-wei Xu, Zong-jie Yao, Yan-di Zhang, Dan-yun Lai, Jo-Lewis Banga Ndzouboukou, Bo Zhang, Hong Chen, Zhu-qing Ouyang, Jun-biao Xue, Xiao-song Lin, Yun-xiao Zheng, Xue-ning Wang, He-wei Jiang, Hai-nan Zhang, Huan Qi, Shu-juan Guo, Mei-an He, Zi-yong Sun, Feng Wang, Sheng-ce Tao, Xiong-lin Fan

## Abstract

The COVID-19 global pandemic is far from ending. There is an urgent need to identify applicable biomarkers for early predicting the outcome of COVID-19. Growing evidences have revealed that SARS-CoV-2 specific antibodies evolved with disease progression and severity in COIVD-19 patients. We assumed that antibodies may serve as biomarkers for predicting disease outcome. By taking advantage of a newly developed SARS-CoV-2 proteome microarray, we surveyed IgG responses against 20 proteins of SARS-CoV-2 in 1,034 hospitalized COVID-19 patients on admission and followed till 66 days. The microarray results were further correlated with clinical information, laboratory test results and patient outcomes. Cox proportional hazards model was used to explore the association between SARS-CoV-2 specific antibodies and COVID-19 mortality. We found that nonsurvivors induced higher levels of IgG responses against most of non-structural proteins than survivors on admission. In particular, the magnitude of IgG antibodies against 8 non-structural proteins (NSP1, NSP4, NSP7, NSP8, NSP9, NSP10, RdRp, and NSP14) and 2 accessory proteins (ORF3b and ORF9b) possessed significant predictive power for patient death, even after further adjustments for demographics, comorbidities, and common laboratory biomarkers for disease severity (all with *p* trend < 0.05). Additionally, IgG responses to all of these 10 non-structural/accessory proteins were also associated with the severity of disease, and differential kinetics and serum positive rate of these IgG responses were confirmed in COVID-19 patients of varying severities within 20 days after symptoms onset. The AUCs for these IgG responses, determined by computational cross-validations, were between 0.62 and 0.71. Our findings have important implications for improving clinical management, and especially for developing medical interventions and vaccines.

## Introduction

The coronavirus disease 2019 (COVID-19), the emerging infectious disease caused by severe acute respiratory syndrome coronavirus 2 (SARS-CoV-2) in December 2019, has quickly become the greatest crisis of global public health, social and economic developments in our times ^1^. As of April 25, 2021, there has been 146.05 million confirmed cases and 3.09 million patients death from SARS-CoV-2 infection worldwide ^2^. Currently, there are no highly effective therapeutics available for the COVID-19 patients ^3-5^. Data from Phase III clinical trials showed that the protective efficacy of vaccines, namely, mRNA-1273 ^6^, BNT162b2 mRNA ^7^, and ChAdOx1 nCoV-19 (AZD1222) ^8^ based on the spike protein of the virus were 94.5%, 95.0% and 70.4%, respectively. However, several vaccinators developed severe allergic symptoms after vaccination ^9,10^, which may belong to vaccine-related immunopathological phenomena though antibody-dependent enhancement (ADE) mechanism ^11,12^. Therefore, it is of significance to elucidate the role of host immune responses in clinical progression and outcome of COVID-19 patients for improving clinical management and developing more effective interventions.

Similar to SARS-CoV and MERS-CoV, SARS-CoV-2 belongs to the beta-coronavirus genus and its genome encodes four major structural proteins, namely, spike (S), envelope (E), membrane (M), and nucleocapsid (N), and 15 non-structural proteins (Nsp1-10 and Nsp12-16), and 8 accessory proteins ^13^. Among these, the S protein, consisted of N-terminal S1 peptide with an important receptor binding domain (RBD) and C-terminal S2 fragment, plays an essential role in viral attachment, fusion, and entry into the target cells which express the viral receptor angiotensin-converting enzyme 2 (ACE2) ^14^. There has been rapidly growing serological evidence that IgM, IgG, and IgA antibodies against S or N proteins of SARS-CoV-2 evolve rapidly in the serum of both asympomatic and symptomatic COVID-19 infections within one week after infection or onset of symptoms ^15-18^. Moreover, these antibodies elevated with disease progression and severity in symptomatic COIVD-19 patients ^19^. Therefore, anti-SARS-CoV-2 specific antibodies may involve in the pathogenesis and affect the disease progression.

In this study, we assumed that levels of anti-SARS-CoV-2 IgG antibodies might help predict the prognosis and outcome of patients with COVID-19. Proteome microarray technology has been confirmed as a mature and repeatable assay, which has been widely used in serological analysis of various diseases ^20-22^. To enable the global understanding of SARS-CoV-2 specific IgG responses and their application, we constructed a proteome microarray with 20 out of the 28 predicted proteins of SARS-CoV-2 ^18,23^. Clinical serum specimens were analyzed on the SARS-CoV-2 proteome microarray, which can provide a high-throughput assay for 12 samples on each microarray and a rapid turnaround time of assay results (within 5 h after sample collection).

1,034 patients hospitalized for confirmed COVID-19 disease at Tongji hospital from the day of hospitalization to the day of discharge or death were enrolled in this study. Serum IgG profiles for 1,034 patients with COVID-19 on admission were probed using the SARS-CoV-2 proteome microarray. The microarray results were further correlated with laboratory biomarkers of disease severity and comorbidities, and with death of each patients, whose known clinical outcomes collected from electronic medical records. We found that the magnitude IgG responses to most of non-structural/accessory proteins were powerful predicting signatures for the COVID-19 death,, independent of other biomarkers of laboratory and clinical severity factors.

## Materials

### Patient information and data source

1,056 confirmed COVID-19 patients were recruited from Tongji Hospital, Wuhan, China, between 17 February 2020 and 28 April 2020. COVID-19 was diagnosed based on positive SARS-CoV-2 nucleic acid test from respiratory tract specimens or based on clinical diagnosis with clinical symptoms and imaging features of pneumonia on chest computed tomographic (CT) according to the fifth version of COVID-19 diagnostic and treatment guideline, published by the National Health Commission of China (NHCC) ^24^. Demographic information, medical history, comorbidities, signs and symptoms, chest CT, laboratory findings during hospitalization, and clinical outcomes were collected from electronic medical records. Among these, laboratory biomarkers related with disease severity factors such as the blood routine (leucocytes, lymphocytes, platelets, and neutrophils), liver and kidney functions (aspartate aminotransferase, alanine aminotransferase, lactate dehydrogenase, and creatinine), coagulation function (D-dimer) and inflammatory biomarkers (C-reactive protein, procalcitonin) were performed by automated analyzers according to the manufacturers’ instructions. The level of IL-2R in serum was measured by an automatic solid-phase two-site chemiluminescent immunometric assay via IMMULITE 1000 Analyzer (Siemens, Germany). Serum IL-6 was measured by an electro-chemiluminescence method (Roche Diagnostics, Switzerland).

Serum specimens were collected from each patient on admission and were stored at -80 °C until use. Serum detection based on proteome microarray and data analysis were performed during April 2020 to March 2021. After excluding 22 individuals whose anti-SARS-CoV-2 antibody indicators were missing more than three, a total of 1,034 eligible participants (524 females and 510 males) with available data from serum proteome microarray and their clinical outcomes were used for the final analysis. Among 1,034 eligible participants, some of whom had serial serum samples and were collected for a total of 2,977 samples.

### Ethical approval

The study was approved by the Ethical Committee of Tongji Hospital, Tongji Medical College, Huazhong University of Science and Technology, Wuhan, China (IRB ID:TJ-C20200128).

### Protein microarray fabrication

The microarray used for serum IgG profiling was prepared as described previously ^18,23^. 20 proteins of SARS-CoV-2 with indicated concentrations, along with the negative (GST, Biotin-control, and eGFP) and positive controls (Human IgG and ACE2-Fc), were printed in quadruplicate on PATH substrate slide (Grace Bio-Labs, USA) to generate identical arrays in a 2×7 subarray format using Super Marathon printer (Arrayjet, UK). The prepared protein microarrays were incubated in blocking buffer (3% BSA in 1×PBS buffer with 0.1% Tween 20) for 3 h, and then stored at -80 °C until use.

### Microarray-based serum analysis

The protein microarrays stored at -80 °C were warmed to room temperature before detection and were performed to probe all available seral samples. A 14-chamber rubber gasket was mounted onto each slide to create individual chambers for the 14 identical subarrays. Serum samples were diluted 1:200 in PBS containing 0.1% Tween 20 and a total of 200 μL of diluted serum or buffer only (negative controls) was incubated with each subarray for 2h at 4°C. The arrays were washed with 1×PBST and bound antibodies were detected by incubating with Cy3-conjugated goat anti-human IgG (Jackson ImmunoResearch, USA), which were diluted 1: 1,000 in 1×PBST, and incubated at room temperature for 1 h. The microarrays were then washed with 1×PBST and dried by centrifugation at room temperature and scanned by LuxScan 10K-A (CapitalBio, China) with the parameters set as 95% laser power/ PMT 480 for IgG. Data of fluorescent intensity (FI) from each microarray was extracted by GenePix Pro 6.0 software (Molecular Devices, USA). The result of FI for each serum response to each protein was defined as the median of the foreground subtracted by the median of background for each spot and then averaged the triplicate spots for each protein. The result of the protein-specific antibody in the serum was expressed as log_2_(FI).

### Statistical analysis

Shapiro-Wilk test was used to test data normality. Two-tailed t-test was conducted to test difference in means between survivor and nonsurvivor groups, Mann-Whitney U test was performed to test difference in skewed parameters. Chi-square tests or Fisher’s exact test, when appropriate, was used for categorical variables. Cox proportional-hazards model was performed to estimate the hazard ratios (HRs) and 95% confidence intervals (CIs) of COVID-19 mortality for individual levels of virus-specific IgG responses categorized into tertiles according to distributions. The lowest tertiles were assigned to be the reference groups. Age and sex were included in Model 1. In Model 2, we further adjusted hypertension (yes/no), diabetes (yes/no), lymphopenia (<1.1, ≥1.1, ×10^9/L), increased alanine aminotransferase (<40, ≥41, U/L), and increased lactate dehydrogenase (<214, ≥214, U/L). Linear trend *p*-values were calculated by modeling the median value of each antibody tertiles as a continuous variable in the adjusted models. Spearman’s rank correlation analysis was performed to explore the correlations between virus-specific IgG responses and laboratory results in the study population. The principal component analysis (PCA) based on the 20 proteins of SARS-CoV-2 specific IgG responses was used to optimize the type of data and extract principal components (PCs). SARS-CoV-2 protein -specific IgG responses with factor loadings over 0.7 on a particular PC were regarded as main contributors of it. Each PC was modeled into the Cox proportional-hazards models as tertiles to evaluate the association with anti-SARS-CoV-2 specific IgG responses and the COVID-19 mortality.

In addition, the results of antibodies were classified as two groups of the high levels (≥ median) and low levels (< median) based on the medians of IgG responses to each protein and further correlated these results with on day 66 mortality of all involved COVID-19 patients by Kaplan-Meier survival curve and log-rank test. Loess regression was used to establish the kinetics of SARS-CoV-2 specific antibodies. Cluster analysis was performed with pheatmap package of R. SAS (version 9.4), R (version 4.0.0), and SPSS (version 23.0) were used to conduct statistical analysis when applicably used. Two-sided statistical tests were considered to be significant at a *p* value below 0.05.

### Computational cross-validations of the prediction efficacy for clinical outcome

The receiver operating characteristic curve was conducted for the prediction of COVID-19 survival and death, and 1,000 times computational cross-validations were conducted. For each cross-validation procedure, 477 survivors and 39 nonsurvivors were randomly selected as the training set. The rest of the samples were treated as the testing set (478 survivors and 40 nonsurvivors).

## Results

### Characteristics of the study population

1,034 participants, having available serum microarray results and consisting of 955 survivors and 79 nonsurvivors, were enrolled in this study. Baseline characteristics of participated patients based on electronic medical records were analyzed as **Table 1**. The median age of all enrolled patients was 63 years old (IQR, 51-71). The median intervals from onset of symptoms to hospital admission, from onset of symptoms to recovery, and from onset of symptoms to death were 13 days (IQR, 8-21), 41 days (IQR, 33-52), and 32 days (IQR, 25-39), respectively. The median length of all COVID-19 patients’ hospital stay was 24 days (IQR, 15-35). 37% patients with COVID-19 had hypertension and 18.5% with diabetes. 30.7% patients had lymphopenia, while increased levels of lactate dehydrogenase and alanine aminotransferase were detected in 43% and 25.4% patients, respectively. Consistent with previous reports ^25,26^, nonsurvivors were more likely to be male, and older than survivors (*p* < 0.001). Higher proportion of abnormal laboratory results and shorter hospitalization time were observed in nonsurvivors than those of survivors (*p* < 0.001).

**Table 1.** Baseline characteristics of participated COVID-19 patients.

|  | All patients | Survivors | Nonsurvivors | *p* value |
| --- | --- | --- | --- | --- |
| N | 1034 | 955 | 79 |  |
| Age, median (IQR), years | 63(51-71) | 62(51-70) | 68(59-78) | <0.001 |
| Female, n (%) | 524(50.7) | 491(51.4) | 33(41.8) | 0.10 |
| Time from onset to admission, Median (IQR), days | 13(8-21) | 13(8-22) | 11(5-19) | 0.03 |
| Length of hospital stay, Median (IQR), days | 24(15-35) | 25(16-35) | 18(9-26) | <0.001 |
| Time from onset to outcome, Median (IQR), days | 40(33-52) | 41(33-52) | 32(25-39) | <0.001 |
| Comorbidity, n (%) |  |  |  |  |
| Hypertension | 383(37.0) | 355(37.2) | 28(35.4) | 0.76 |
| Diabetes | 191(18.5) | 173(18.1) | 18(22.8) | 0.30 |
| Coronary heart disease | 68(6.6) | 57(6.0) | 11(13.9) | 0.006 |
| Chronic obstructive pulmonary disease | 6(0.6) | 3(0.3) | 3(3.8) | 0.007 |
| Cerebrovascular disease | 44(4.3) | 37(3.9) | 7(8.9) | 0.07 |
| Chronic liver disease | 21(2.0) | 19(2.0) | 2(2.5) | 0.67 |
| Chronic renal disease | 23(2.2) | 20(2.1) | 3(3.8) | 0.41 |
| Cancer | 45(4.4) | 35(3.7) | 10(12.7) | 0.001 |
| Laboratory results, n (%) |  |  |  |  |
| Lymphopenia, $<1.1 \times 10^9/L$ | 294(30.7) | 234(26.4) | 60(83.3) | <0.001 |
| Neutrophilia, $\geq 6.3 \times 10^9/L$ | 181(18.9) | 125(14.1) | 56(77.8) | <0.001 |
| Thrombocytopenia, $\geq 350 \times 10^9/L$ | 64(6.7) | 62(7.0) | 2(2.7) | 0.16 |
| Leukocytosis, $\geq 9.5 \times 10^9/L$ | 146(15.2) | 98(11.1) | 48(65.8) | <0.001 |
| Increased lactate dehydrogenase, $\geq 214$ U/L | 405(43.0) | 342(39.3) | 63(88.7) | <0.001 |
| Increased alanine aminotransferase, $\geq 41$ U/L | 239(25.4) | 217(24.9) | 22(31.0) | 0.26 |
| Increased aspartate aminotransferase, $\geq 40$ U/L | 129(13.7) | 101(11.6) | 28(40.0) | <0.001 |
| Increased creatinine, $\geq 104$ $\mu\text{mol/L}$ | 57(6.3) | 39(4.7) | 18(26.1) | <0.001 |
| Increased C-reactive protein, $\geq 3\text{mg/L}$ | 330(45.9) | 289(42.7) | 41(97.6) | <0.001 |
| Increased procalcitonin, $\geq 0.05$ ng/ml | 159(29.3) | 122(24.3) | 37(92.5) | <0.001 |
| Increased D-dimer, $\geq 0.5$ mg/L | 361(59.4) | 302(55.1) | 59(98.3) | <0.001 |
| Increased IL2R, $>710$ U/mL | 67(16.2) | 57(14.4) | 10(55.6) | <0.001 |
| Increased IL6, $>7$ ng/L | 98(23.5) | 82(20.6) | 16(88.9) | <0.001 |

Data were shown as medians (IQR) or number (%), respectively. IQR, inter-quartile ranges.

### Nonsurvivors produce higher levels of IgG responses against most of non-structural proteins than survivors

To establish the association of anti-SARS-CoV-2 IgG antibodies with COVID-19 survival and death, serum collected from each involved patients on admission was used for microarray-based serum analysis. Based on the FI value extracted from the proteome microarray for each serum sample of 1034 patients, we first compared IgG profiles against 20 proteins of SARS-CoV-2 (**Table 2**). There was no statistical difference of the levels of either anti-S or N IgG antibodies between nonsurvivors and survivors. However, higher levels of IgG responses against 15 proteins, namely, E, NSP1, NSP2, NSP4, NSP5, NSP7, NSP8, NSP9, NSP10, RdRp, NSP14, NSP15, NSP16, ORF3b and ORF9b, were induced in nonsurvivors than those of survivors. Our results indicate that the magnitude of IgG responses against most of non-structural proteins of SARS-CoV-2 might predict the prognosis and outcome of COVID-19.

**Table 2.**
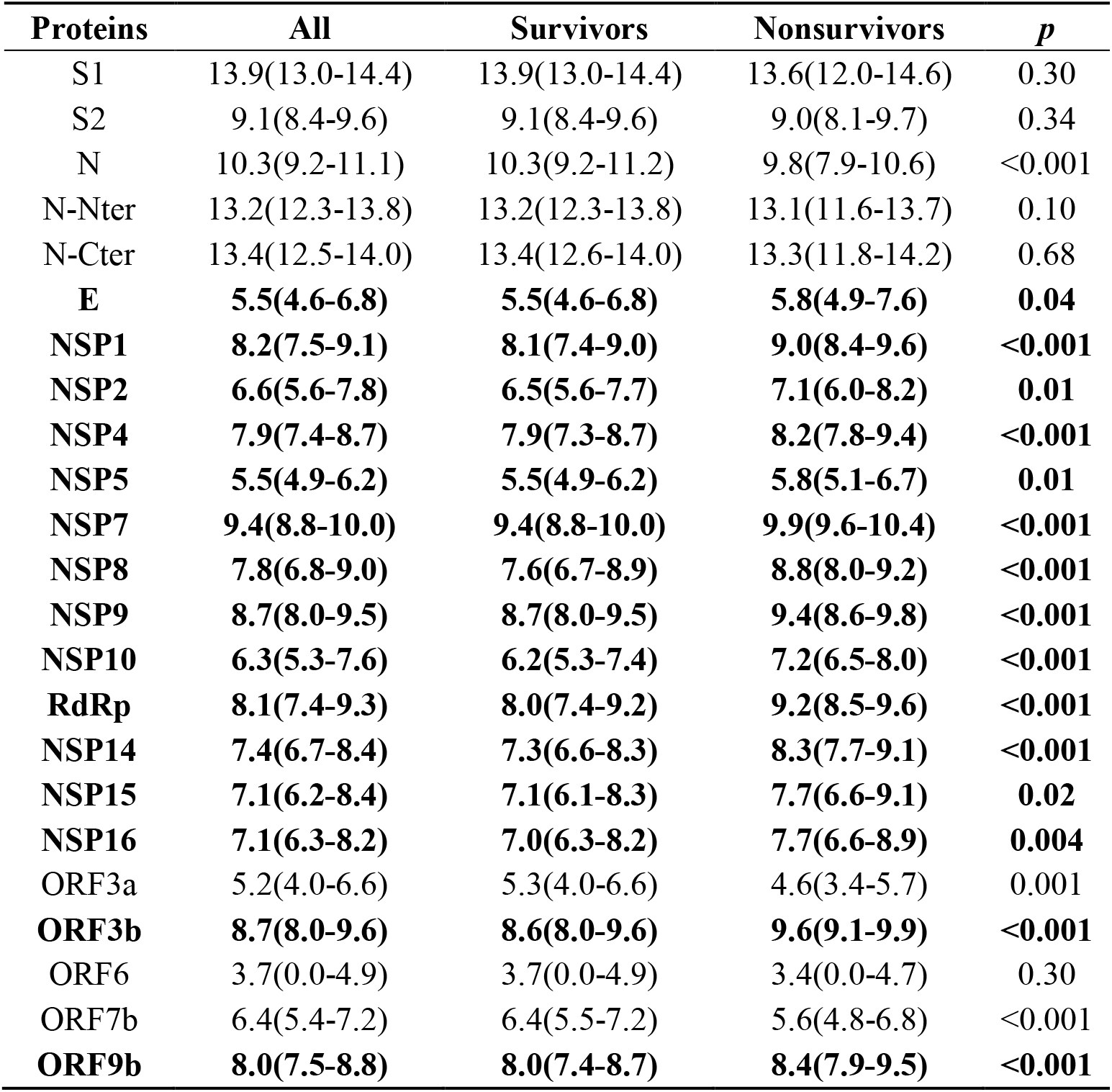
Comparison of SARS-CoV-2 specific IgG responses ([log2(FI)]) between survivors and nonsurivivors.

| **Proteins** | **All** | **Survivors** | **Nonsurvivors** | ***p*** |
| --- | --- | --- | --- | --- |
| S1 | 13.9(13.0-14.4) | 13.9(13.0-14.4) | 13.6(12.0-14.6) | 0.30 |
| S2 | 9.1(8.4-9.6) | 9.1(8.4-9.6) | 9.0(8.1-9.7) | 0.34 |
| N | 10.3(9.2-11.1) | 10.3(9.2-11.2) | 9.8(7.9-10.6) | <0.001 |
| N-Nter | 13.2(12.3-13.8) | 13.2(12.3-13.8) | 13.1(11.6-13.7) | 0.10 |
| N-Cter | 13.4(12.5-14.0) | 13.4(12.6-14.0) | 13.3(11.8-14.2) | 0.68 |
| **E** | **5.5(4.6-6.8)** | **5.5(4.6-6.8)** | **5.8(4.9-7.6)** | **0.04** |
| **NSP1** | **8.2(7.5-9.1)** | **8.1(7.4-9.0)** | **9.0(8.4-9.6)** | **<0.001** |
| **NSP2** | **6.6(5.6-7.8)** | **6.5(5.6-7.7)** | **7.1(6.0-8.2)** | **0.01** |
| **NSP4** | **7.9(7.4-8.7)** | **7.9(7.3-8.7)** | **8.2(7.8-9.4)** | **<0.001** |
| **NSP5** | **5.5(4.9-6.2)** | **5.5(4.9-6.2)** | **5.8(5.1-6.7)** | **0.01** |
| **NSP7** | **9.4(8.8-10.0)** | **9.4(8.8-10.0)** | **9.9(9.6-10.4)** | **<0.001** |
| **NSP8** | **7.8(6.8-9.0)** | **7.6(6.7-8.9)** | **8.8(8.0-9.2)** | **<0.001** |
| **NSP9** | **8.7(8.0-9.5)** | **8.7(8.0-9.5)** | **9.4(8.6-9.8)** | **<0.001** |
| **NSP10** | **6.3(5.3-7.6)** | **6.2(5.3-7.4)** | **7.2(6.5-8.0)** | **<0.001** |
| **RdRp** | **8.1(7.4-9.3)** | **8.0(7.4-9.2)** | **9.2(8.5-9.6)** | **<0.001** |
| **NSP14** | **7.4(6.7-8.4)** | **7.3(6.6-8.3)** | **8.3(7.7-9.1)** | **<0.001** |
| **NSP15** | **7.1(6.2-8.4)** | **7.1(6.1-8.3)** | **7.7(6.6-9.1)** | **0.02** |
| **NSP16** | **7.1(6.3-8.2)** | **7.0(6.3-8.2)** | **7.7(6.6-8.9)** | **0.004** |
| ORF3a | 5.2(4.0-6.6) | 5.3(4.0-6.6) | 4.6(3.4-5.7) | 0.001 |
| **ORF3b** | **8.7(8.0-9.6)** | **8.6(8.0-9.6)** | **9.6(9.1-9.9)** | **<0.001** |
| ORF6 | 3.7(0.0-4.9) | 3.7(0.0-4.9) | 3.4(0.0-4.7) | 0.30 |
| ORF7b | 6.4(5.4-7.2) | 6.4(5.5-7.2) | 5.6(4.8-6.8) | <0.001 |
| **ORF9b** | **8.0(7.5-8.8)** | **8.0(7.4-8.7)** | **8.4(7.9-9.5)** | **<0.001** |

### IgG responses against 10 non-structural/accessory proteins positively correlate with COVID-19 mortality risk

To assess the relationship of the magnitude of IgG antibodies with the mortality risk of COVID-19 patients, the HRs (95% CIs) for the mortality risk associated with the levels of IgG responses against different proteins of SARS-CoV-2 were categorized into tertiles (**Table 3**). We first analyzed the effects of age and gender on the disease death as model 1. After adjusting for age and gender, we found that IgG responses to 10 proteins (NSP1, NSP4, NSP7, NSP8, NSP9, NSP10, RdRp, NSP14, ORF3b and ORF9b) were significantly positively associated with the COVID-19 mortality, whereas negative significant association was observed between N, ORF3a, and ORF7b-specific IgG responses and the death. Previous studies reported that comorbidities and laboratory biomarkers related with the function of important organs also might be the risk factors of the COVID-19 death ^26,27^. Therefore, we further adjusted the association for hypertension, diabetes, lymphopenia, increased alanine aminotransferase and lactate dehydrogenase as shown in model 2. Interestingly, IgG responses to 10 proteins (NSP1, NSP4, NSP7, NSP8, NSP9, NSP10, RdRp, NSP14, ORF3b and ORF9b) were also significantly positively associated with the mortality risk of COVID-19 (**Table 3**).

**Table 3.**
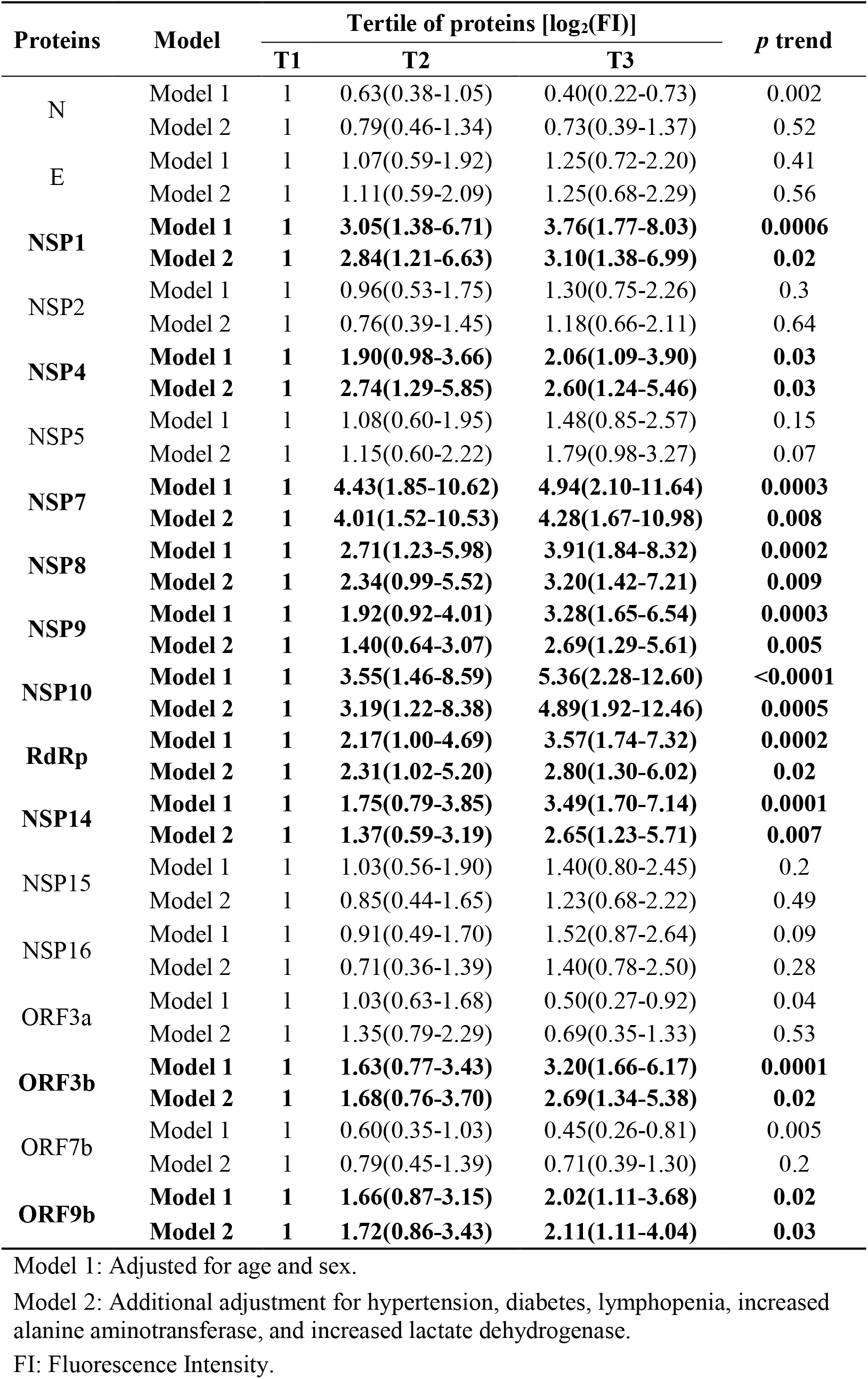
Hazard ratio (95%CI) for COVID-19 mortality according to tertiles of anti-SARS-CoV-2 specific IgG responses.

The Kaplan-Meier survival curve also supported that COVID-19 patients with higher levels of specific IgG responses against NSP1 (log_2_FI ≥ 8.2), NSP4 (log_2_FI ≥ 7.9), NSP7 (log_2_FI ≥ 9.4), NSP8 (log_2_FI ≥ 7.8), NSP9 (log_2_FI ≥ 8.7), NSP10 (log_2_FI ≥ 6.3), RdRp (log_2_FI ≥ 8.1), NSP14 (log_2_FI ≥ 7.4), ORF3b (log_2_FI ≥ 8.7), and ORF9b (log_2_FI ≥ 8.0) had higher morality risk after admission, respectively (**Figure 1**).

**Figure 1.**
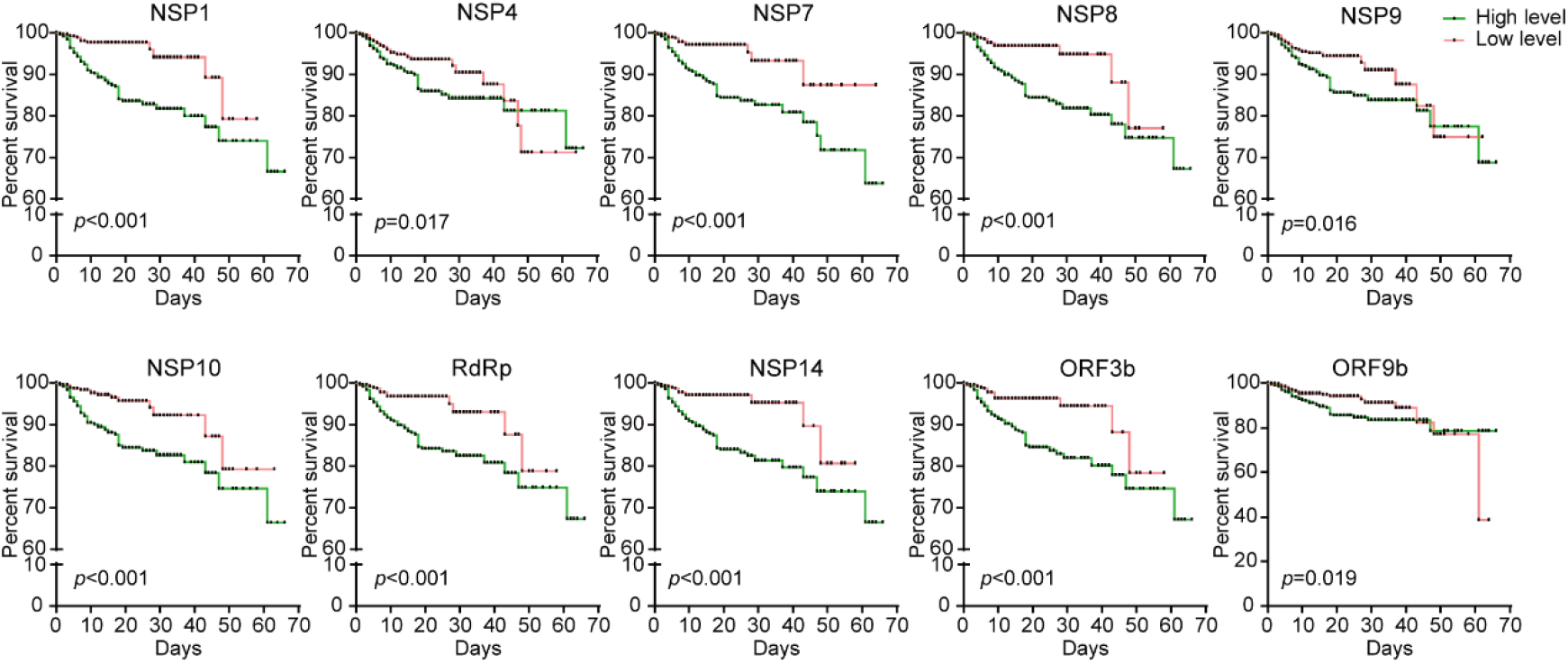
Kaplan-Meier survival curves of patients with high and low levels of IgG to 10 non-structural/accessory proteins. Based on the median level of IgG responses to each protein, patients were classified as both high and low level groups after admission. Kaplan-Meier survival curves of patients with high (green) and low (red) levels of IgG antibodies to each protein, and Log-rank test was used to analyze the difference between two groups.

To further establish the association among IgG responses to different proteins with the outcome of COVID-19, we further conducted principal component analyses (PCs) and screened hypothetical new variables that account for the variance as much as possible, in order to reduce the dimension of data and the complexity of data with the least loss of original information. The HRs (95%CIs) for the COVID-19 mortality according to PCs tertiles were presented in **Table 4**. Four PCs with eigenvalues > 1 were extracted, accounting for 71.95% of the total variance. Of four PCs, we found that only PC1 had the statistical association with the COVID-19 mortality (*p* trend = 0.004, **Table 4**), whatever adjusting age and sex, or further for hypertension, diabetes, lymphopenia, increased alanine aminotransferase and lactate dehydrogenase. Interestingly, IgG responses to 10 proteins (NSP1, NSP4, NSP7, NSP8, NSP9, NSP10, RdRp, NSP14, ORF3b and ORF9b) remained main contributors of PC1 (**Table 5**), in line with our above findings.

**Table 4.**
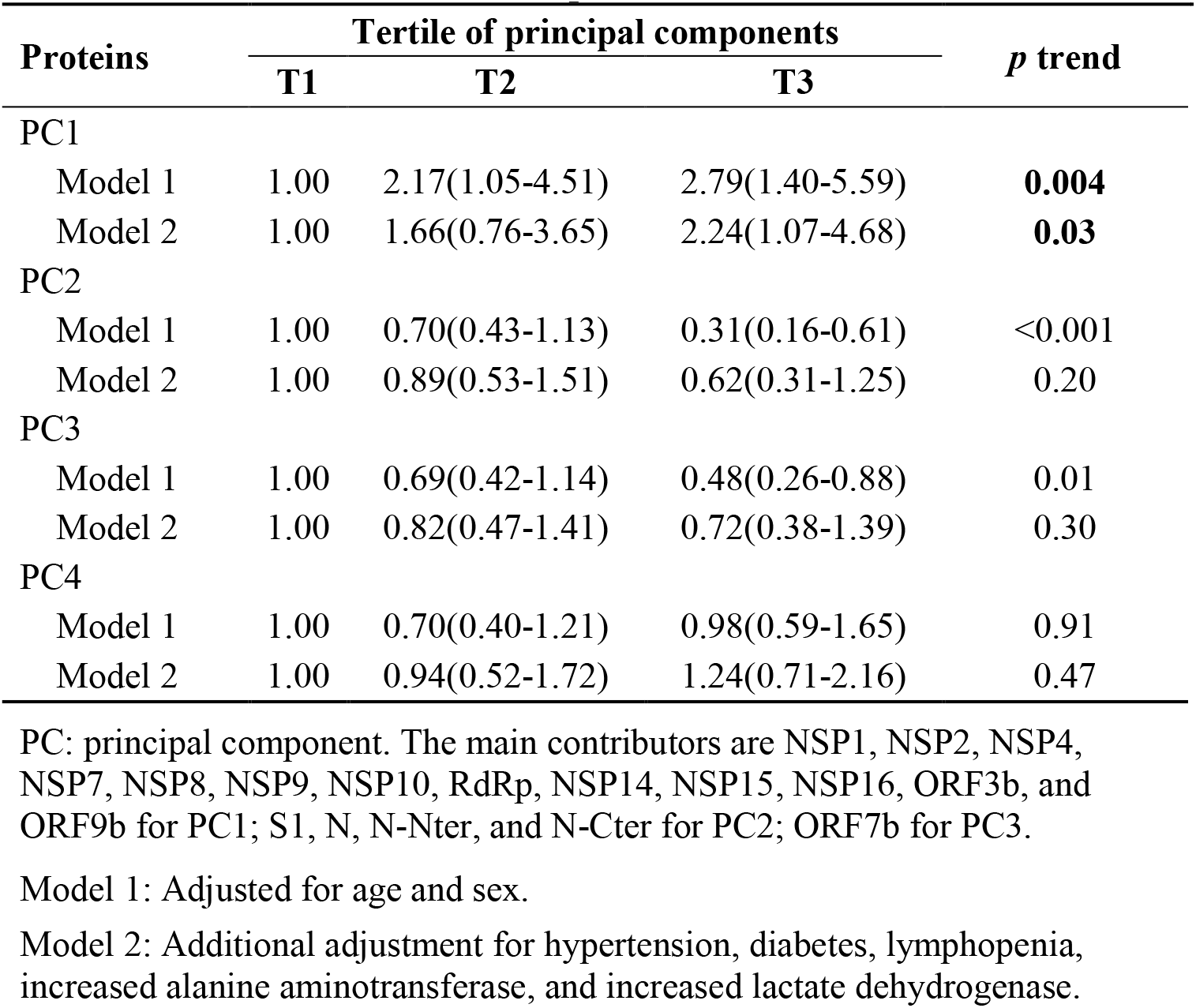
Hazard ratio (95%CI) for COVID-19 mortality according to tertiles of principal components of anti-SARS-CoV-2 specific IgG responses.

**Table 5.** Factor loadings of 20 proteins of anti-SARS-CoV-2 specific IgG responses among the study participants.

| Variables | PC1 | PC2 | PC3 | PC4 |
| --- | --- | --- | --- | --- |
| S1 | 0.26 | **0.87** | -0.15 | -0.06 |
| S2 | 0.36 | 0.59 | -0.05 | 0.27 |
| N | 0.15 | **0.87** | 0.07 | -0.07 |
| N-Nter | 0.26 | **0.90** | -0.08 | -0.07 |
| N-Cter | 0.39 | **0.83** | -0.12 | -0.08 |
| E | 0.67 | -0.07 | 0.45 | -0.29 |
| NSP1 | **0.87** | -0.13 | -0.13 | 0.09 |
| NSP2 | **0.78** | -0.04 | 0.27 | -0.17 |
| NSP4 | **0.87** | -0.10 | -0.06 | 0.11 |
| NSP5 | 0.65 | 0.01 | 0.49 | -0.12 |
| NSP7 | **0.78** | -0.05 | -0.18 | 0.14 |
| NSP8 | **0.79** | -0.16 | -0.21 | 0.13 |
| NSP9 | **0.72** | -0.13 | -0.22 | 0.25 |
| NSP10 | **0.77** | -0.19 | -0.28 | 0.19 |
| RdRp | **0.81** | -0.12 | -0.21 | 0.00 |
| NSP14 | **0.89** | -0.05 | 0.07 | -0.20 |
| NSP15 | **0.78** | -0.09 | 0.15 | -0.17 |
| NSP16 | **0.81** | -0.04 | 0.29 | -0.21 |
| ORF3a | -0.20 | 0.29 | 0.50 | 0.47 |
| ORF3b | **0.85** | -0.13 | -0.13 | 0.09 |
| ORF6 | 0.17 | 0.04 | 0.23 | 0.67 |
| ORF7b | 0.18 | -0.02 | **0.72** | 0.19 |
| ORF9b | **0.78** | -0.05 | -0.10 | 0.03 |
| Eigen values | 9.95 | 3.609 | 1.79 | 1.198 |
| Total variance (%) | 43.263 | 15.691 | 7.784 | 5.207 |
| Cumulative variance (%) | 43.263 | 58.954 | 66.737 | 71.945 |

Bold values denote factor loading > 0.7 are deemed to be statistically significant.

In addition, previous studies have established the associations between COVID-19 death with several laboratory measurements, such as lymphocyte count, procalcitonin, C-reactive protein, lactate dehydrogenase, D-dimer, IL-2R, and IL-6 ^25-27^. Linear correlation between SARS-CoV-2 specific IgG responses with these biomarkers was further analyzed (**Table 6**). Interestingly, IgG responses to 10 proteins were positively correlated with most of these biomarkers but negatively associated with the lymphocyte count. Taken together, our results confirmed that IgG responses to 10 non-structural/accessory proteins were positively correlated with the mortality risk of COVID-19.

**Table 6.** Correlations between the levels of anti-SARS-CoV-2 specific IgG responses and other laboratory biomarkers related with severity factors.

|  | PCT | CRP | LYMPH | LDH | DD | IL-2R | IL-6 |
| --- | --- | --- | --- | --- | --- | --- | --- |
| NSP1_IgG |  |  |  |  |  |  |  |
| $r_s$ | 0.19** | 0.21** | -0.16** | 0.17** | 0.31** | 0.18** | 0.09 |
| NSP4_IgG |  |  |  |  |  |  |  |
| $r_s$ | 0.09* | 0.14** | -0.09** | 0.10** | 0.21** | 0.10* | 0.02 |
| NSP7_IgG |  |  |  |  |  |  |  |
| $r_s$ | 0.19** | 0.22** | -0.17** | 0.19** | 0.31** | 0.14** | 0.08 |
| NSP8_IgG |  |  |  |  |  |  |  |
| $r_s$ | 0.12** | 0.19** | -0.15** | 0.16** | 0.31** | 0.11* | 0.12* |
| NSP9_IgG |  |  |  |  |  |  |  |
| $r_s$ | 0.12** | 0.17** | -0.09** | 0.12** | 0.17** | 0.07 | 0.07 |
| NSP10_IgG |  |  |  |  |  |  |  |
| $r_s$ | 0.12** | 0.21** | -0.15** | 0.15** | 0.31** | 0.16** | 0.13** |
| RdRp_IgG |  |  |  |  |  |  |  |
| $r_s$ | 0.17** | 0.19** | -0.15** | 0.14** | 0.31** | 0.13** | 0.11* |
| NSP14_IgG |  |  |  |  |  |  |  |
| $r_s$ | 0.15** | 0.17** | -0.15** | 0.16** | 0.27** | 0.17** | 0.11* |
| ORF3b_IgG |  |  |  |  |  |  |  |
| $r_s$ | 0.15** | 0.18** | -0.14** | 0.16** | 0.29** | 0.12* | 0.06 |
| ORF9b_IgG |  |  |  |  |  |  |  |
| $r_s$ | 0.12** | 0.12** | -0.07* | 0.11** | 0.19** | 0.04 | 0.01 |

Spearman's rank correlation coefficients were shown in the table. \* $p < 0.05$ , \*\* $p < 0.01$ . PCT: procalcitonin; CRP: C-reactive protein; LYMPH: lymphocyte count; LDH: lactate dehydrogenase; DD: D-dimer; IL-2R: interleukin-2 receptor; IL-6: interleukin-6.

### IgG responses against 10 non-structural/accessory proteins are associated with the severity of COVID-19 disease

To assess the role of IgG to 10 non-structural/accessory proteins for the prediction of the clinical outcome, signal intensities and serum positive rates of IgG antibodies against 10 non-structural/accessory proteins in 1,034 COVID-19 patients were compared with those of 601 healthy human serum controls. The cut-off value was set as mean + 2SD of the control group, and positive rates was calculated for each protein. Interestingly, COVID-19 patients had stronger signal intensities of serum IgG responses to all of these 10 proteins than healthy controls (**Figure 2**). In addition, the serum positive rates of IgG antibodies in COVID-19 patients ranged from 7.0% to 50.6%, varying with different proteins. ORF3b, NSP7, and NSP1 specific IgG antibodies listed the top three of the serum positive rates in COVID-19 patients (**Figure 2**).

**Figure 2.**
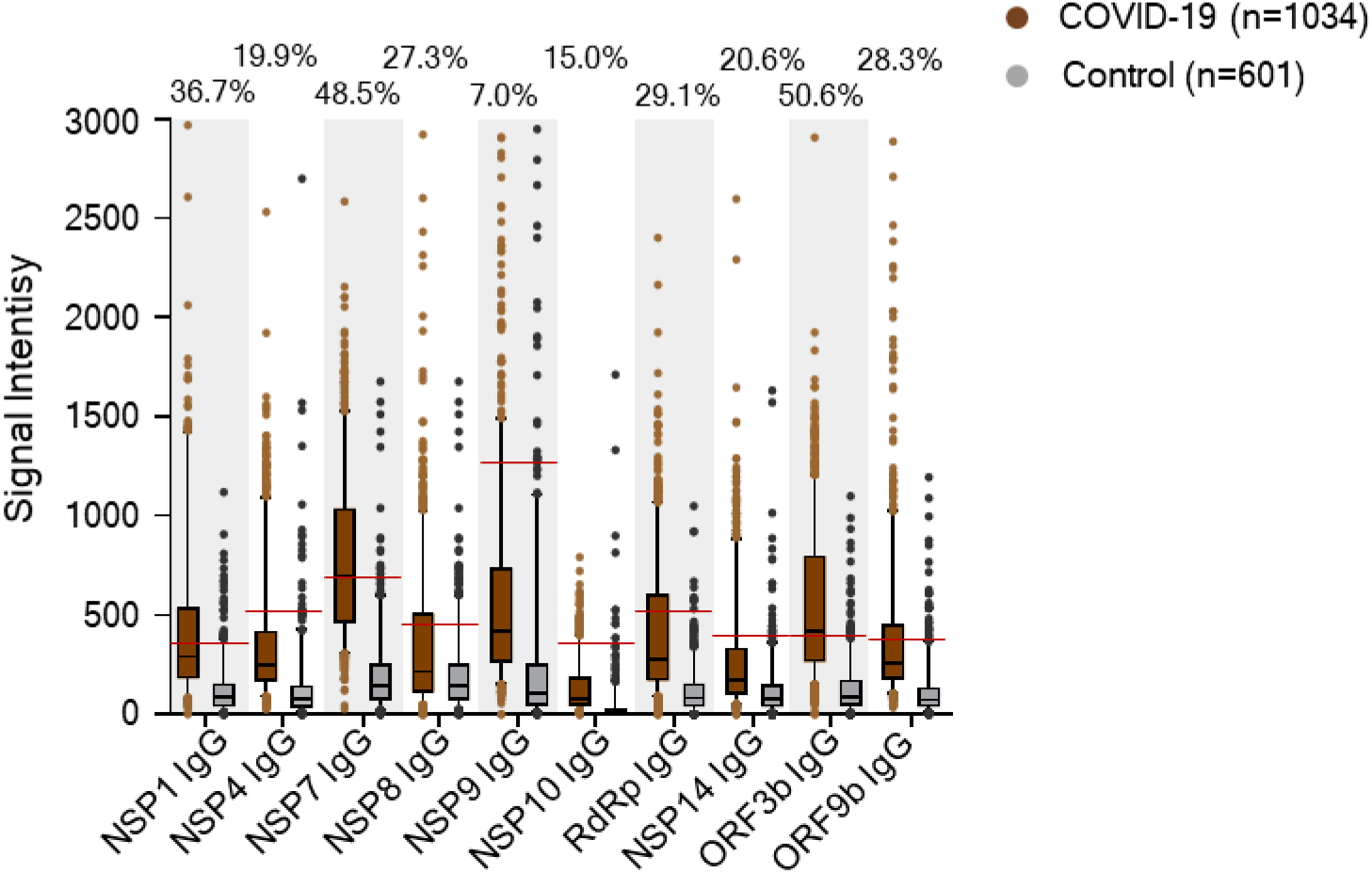
Comparison of signal intensities and positive rates of IgG antibodies between COVID-19 patients and healthy controls. IgG responses to 10 non-structural/accessory proteins were compared between 1,034 COVID-19 patients and 601 healthy serum controls. IgG responses were depicted as the boxplot according to the signal intensity of each serum sample on the proteome microarray. Data were represented by the median and 5th-95th percentile. The cut-off values of IgG antibody to each protein were set as mean + 2SD of the control group (n = 601) and shown as the red line. The positive rates of IgG antibodies to each protein in the patient groups were labeled on the figure.

To further explore the association of IgG antibodies with the severity of illness, 1,034 COVID-19 patients included in this study were divided into three groups: non-severe (n=508), severe-survivors (n=447), and severe-nonsurvivors (n=79). Both the serum positive rate and the signal intensity of IgG responses were compared among these groups (**Figure 3**). Interestingly, severe-nonsurvivors had higher serum positive rates of NSP1, NSP7, NSP8, RdRp, ORF3b and ORF9b specific IgG antibodies than severe-survivors and the non-severe group. In addition, the overall signal intensities for the 10 protein-specific IgG antibodies were higher in severe-nonsurvivors than those of severe-survivors (**Figure 3**). These results suggested that the IgG responses of 10 non-structural/accessory proteins were also associated with the disease severity and might be effective predictors of disease prognosis.

**Figure 3.**
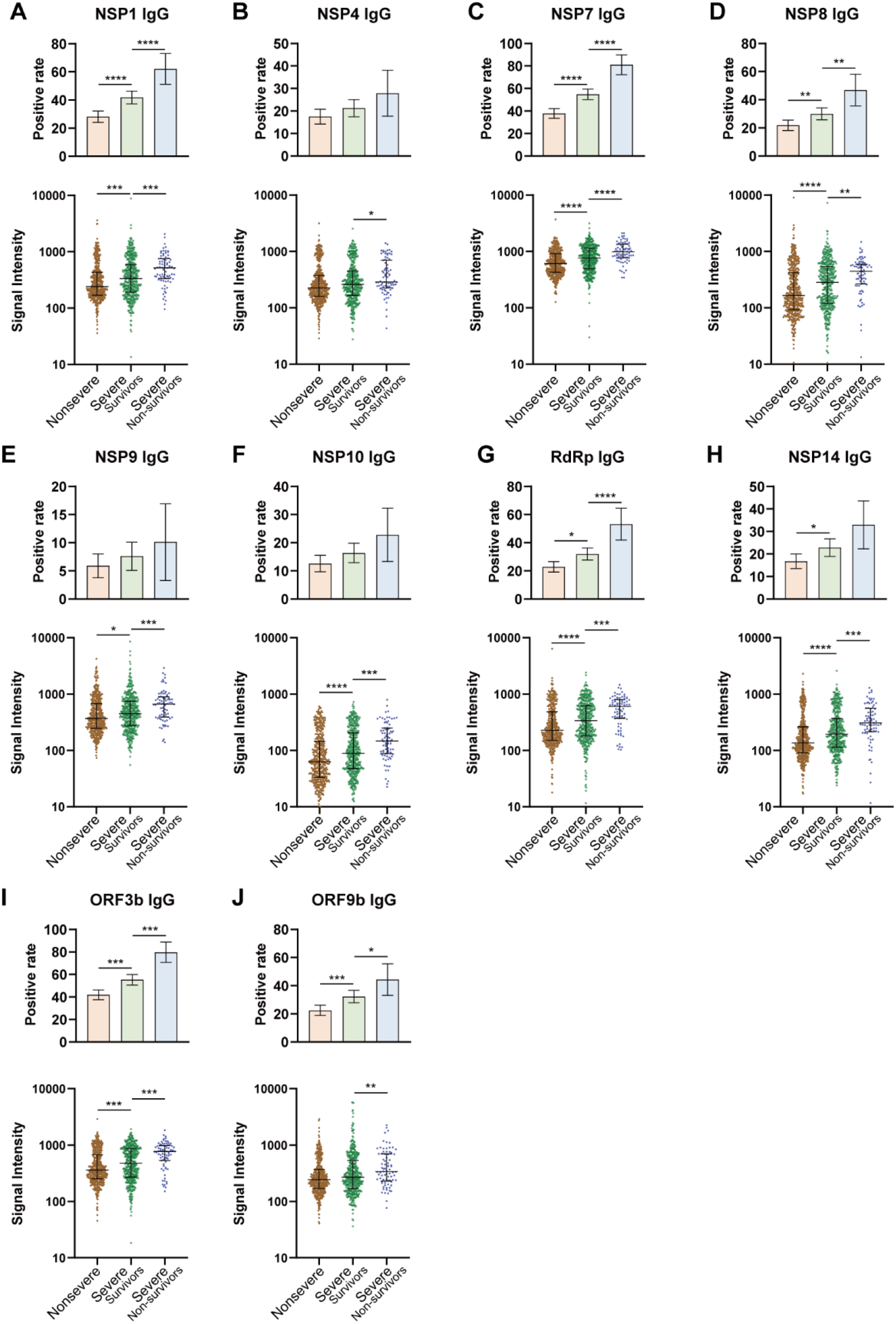
Comparison of IgG responses of 10 non-structural/accessory proteins among different severities of patients. 1,034 COVID-19 patients included in this study were divided into three groups: non-severe (n=508), severe-survivors (n=447), and severe-nonsurvivors (n=79). Serum positive rate and signal intensity of IgG responses to NSP1 (A), NSP4 (B), NSP7 (C), NSP8 (D), NSP9 (E), NSP10 (F), RdRp (G), NSP14 (H), ORF3b (I), and ORF9b (J) were compared among different groups. For the positive rate analysis, error bar was given as the 95% confidential interval, and χ^2^ test was used to calculate *p* values. For the signal intensity analysis, the middle line was set as the median value; the upper and lower hinges were the values of 75% and 25% percentile, and Kruskale Wallis test and post-hoc test (Dunn-Bonferroni) were conducted to calculate *p* values. \**p* <0.05,\*\**p* <0.01,\*\*\**p* <0.001,\*\*\*\**p* <0.0001.

### IgG responses to 10 non-structural/accessory proteins peak within 20 days after onset

To explore the detection time of IgG responses for the prediction, we further established the dynamic of IgG responses to 10 non-structural/accessory proteins from 0 to 60 days after onset, using 2,977 seral samples from 1,034 COVID-19 patients. Overall, the signal intensity and serum positive rate of the 10 protein-specific IgG antibodies increased persistently with the time after the symptom onset, peaked about 20 days later, and then declined gradually (**Figure 4**). Interestingly, severe-nonsurvivors had a stronger signal intensity and higher serum positive rate than non-severe and severe-survivors. Our results indicated that detection of these antibodies within 20 days after the symptom onset might be used to predict the prognosis of disease.

**Figure 4.**
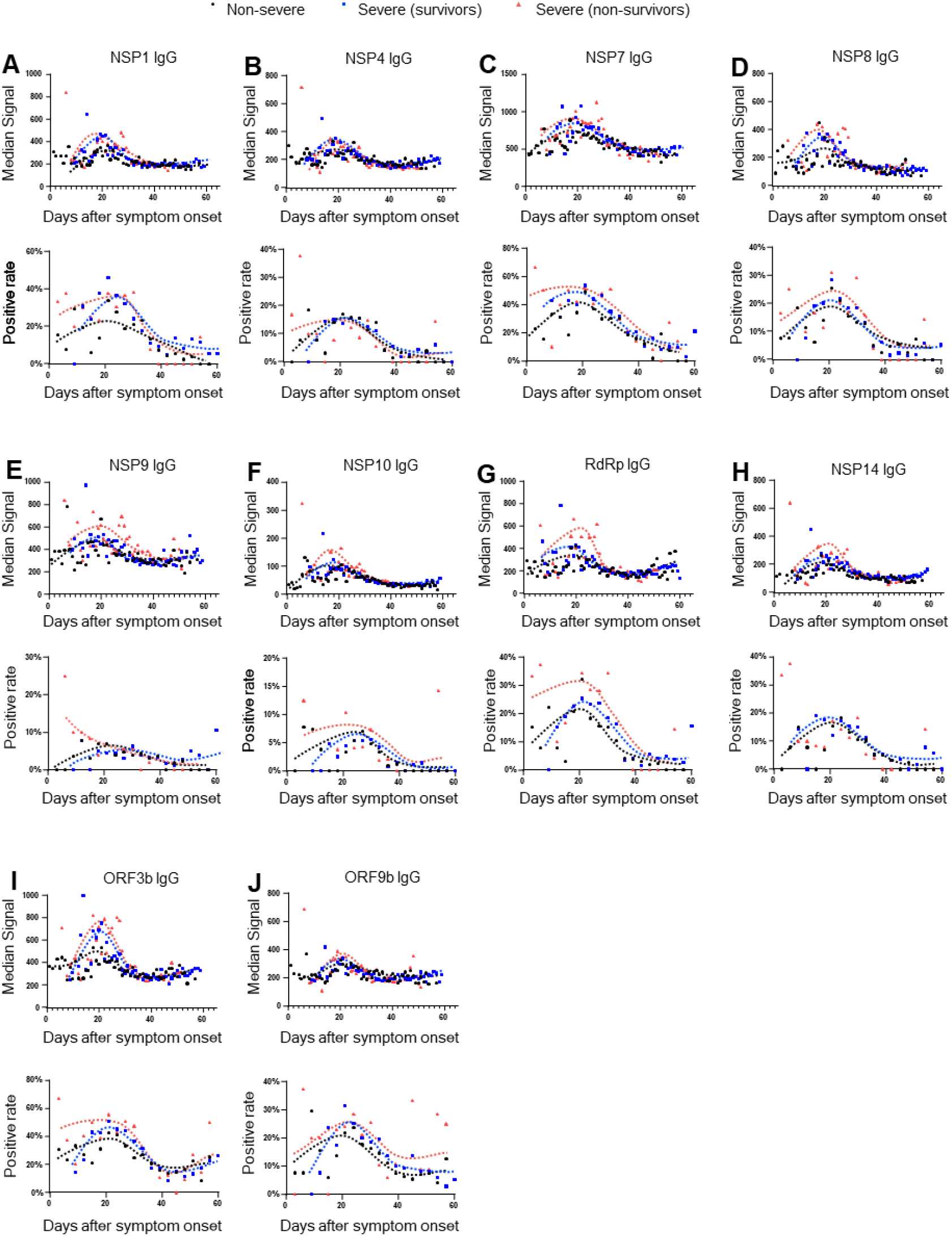
The dynamics of IgG responses to 10 non-structural/accessory proteins between different groups. 2,977 seral samples from 1,034 COVID-19 patients were used. The patients were divided into three groups: non-severe (n=508), severe-survivors (n=447), and severe-nonsurvivors (n=79). Signal intensity and serum positive rate of IgG responses to NSP1 (A), NSP4 (B), NSP7 (C), NSP8 (D), NSP9 (E), NSP10 (F), RdRp (G), NSP14 (H), ORF3b (I), and ORF9b (J) were compared among different groups. For signal intensity analysis, samples were grouped per day and the points with sample number less than 4 were excluded. For positive rate analysis, samples were grouped per three days.

### Validation models confirm high prediction efficacy of IgG antibodies for clinical outcome

It is a common practice to validate “potential biomarker” by independent sample cohort. However, it is very difficult to collect new COVID-19 serum samples in China. To assure the reliability of our finding, we performed computational cross-validation based on the large sample cohort, by following protocols as established previously ^21^. IgG response to proteins (NSP1, NSP4, NSP7, NSP8, NSP9, NSP10, RdRp, NSP14, ORF3b and ORF9b) were explored as 10 potential biomarkers for predicting clinical outcome as shown in **Figure 5**. Interestingly, the AUCs of these IgG antibodies for predicting COVID-19 death ranged from 0.62 and 0.71 (**Figure 6**). NSP7, RdRp, and NSP14 specific IgG listed the top three of high AUC values.

**Figure 5.**
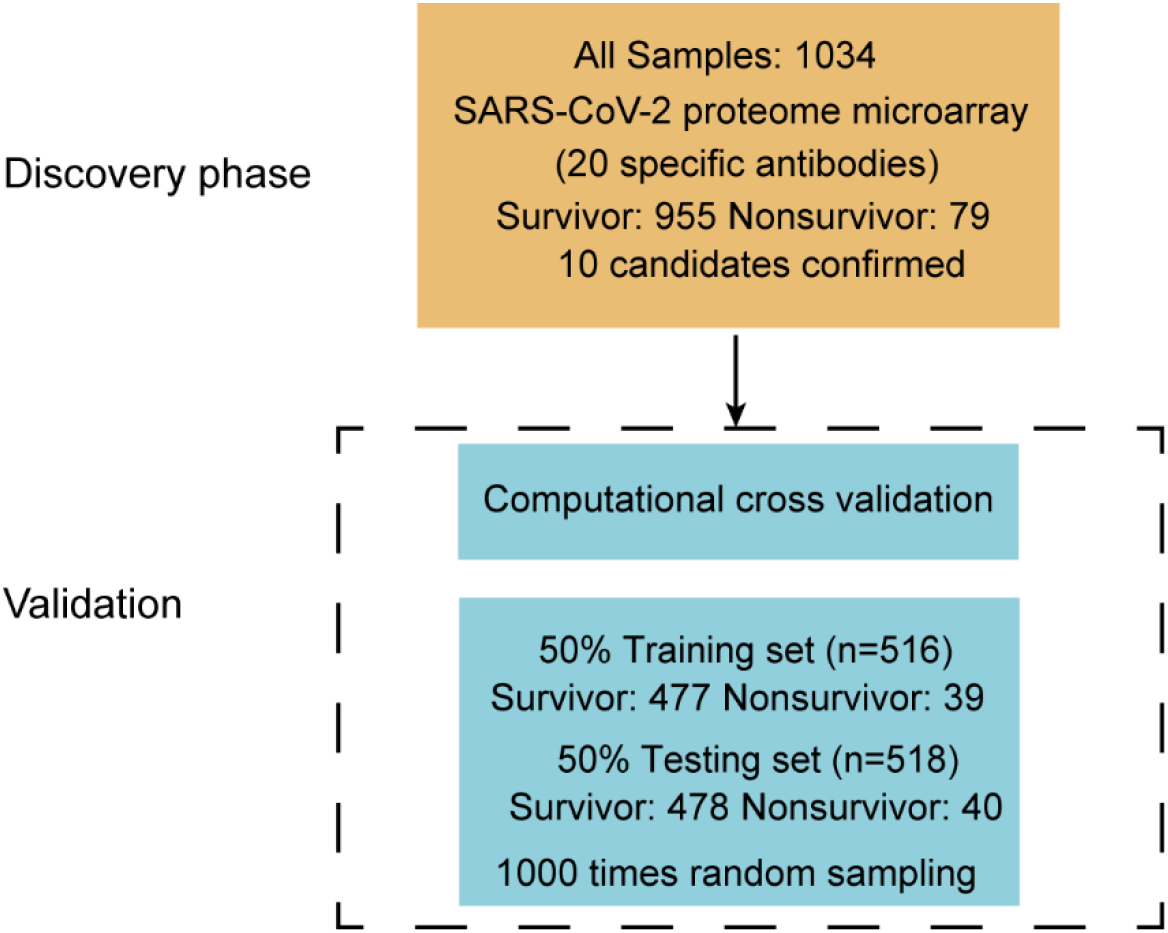
The workflow of validation.

**Figure 6.**
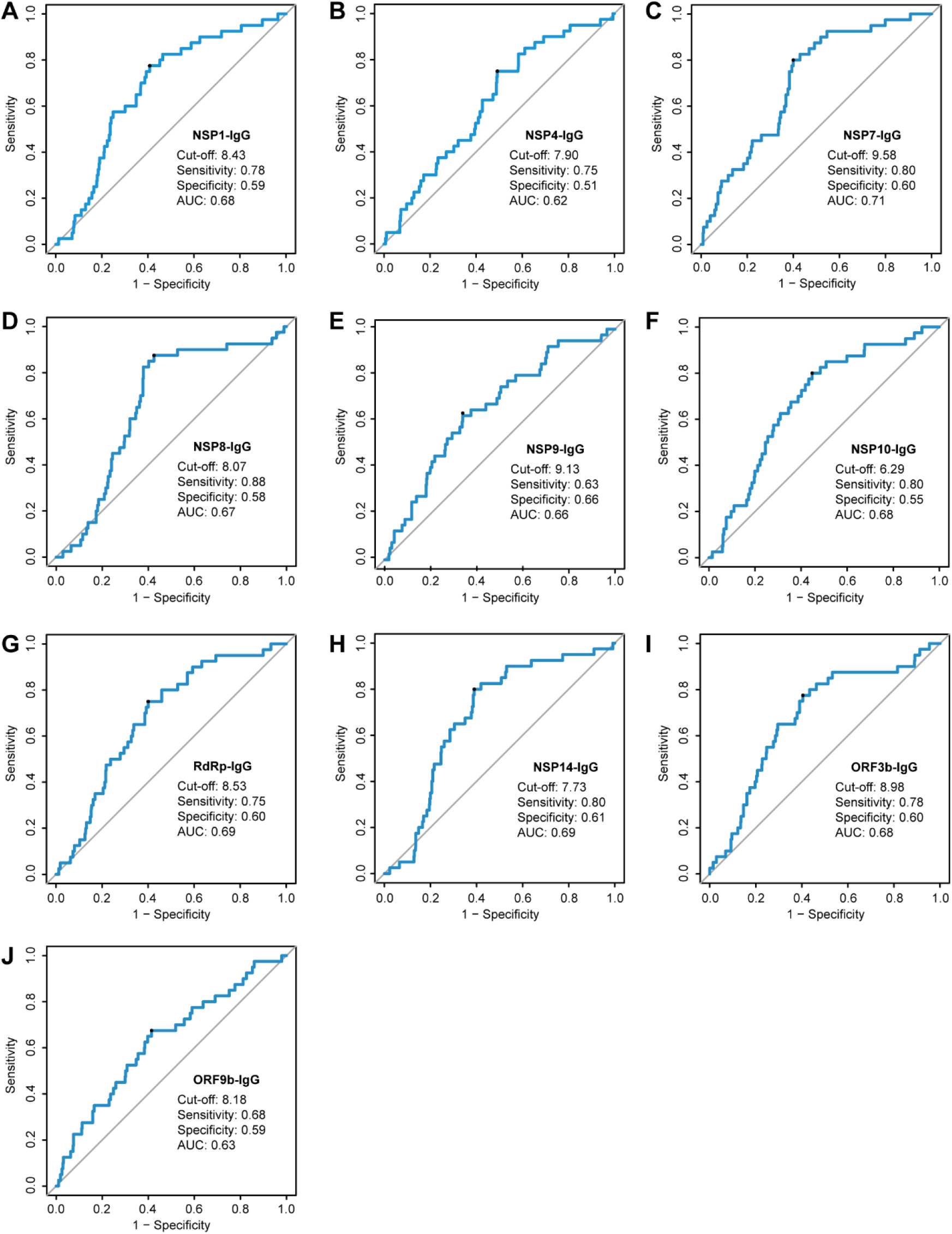
Computational cross-validations of IgG responses to 10 non-structural/accessory proteins for the prediction efficacy. The prediction efficacy was determined by a computational cross-validation. The receiver operating characteristic curve was conducted for the prediction of COVID-19 survival and death, and 1,000 times computational cross-validations were conducted. For each cross-validation procedure, 477 survivors and 39 non-survivors were randomly selected as the training set. The rest of the samples were treated as the testing set (478 survivors and 40 non-survivors). The average cutoff values were shown.

## Discussion

In this study, we demonstrated that IgG responses to 10 non-structural/accessory, namely, NSP1, NSP4, NSP7, NSP8, NSP9, NSP10, RdRp, NSP14, ORF3b, and ORF9b of SARS-CoV-2 were significantly positively associated with the mortality risk and disease severity of COVID-19, which are powerful predicting signatures for predicting clinical outcome. Our findings have important indications for medical interventions and better control of the COVID-19 pandemic.

Firstly, we established a rapid and high-throughput assay platform based on proteome microarrays to measure IgG responses against 20 SARS-CoV-2 proteins in the serum of COVID-19 patients. After analyzing 1,034 hospitalized patients, we found that the outcome of COVID-19 is associated with high levels of IgG responses to 10 non-structural/accessory proteins of SARS-CoV-2 at presentation. Importantly, our observations indicated that antibody patterns are predictive of COVID-19 mortality, independently of demographics and comorbidities, as well as routine clinical biomarkers of disease severity. In particular, we found that IgG antibodies against 8 non-structural proteins (NSP1, NSP4, NSP7, NSP8, NSP9, NSP10, RdRp, and NSP14) and 2 accessory proteins (ORF3b and ORF9b) were predictors of death after adjusting for the demographic features and comorbidities. Early IgG antibody measurements based on our established serum proteome microarray analysis as predictors of mortality, therefore, raise the importance of using antibody levels for rapidly improving clinical management, treatment decisions and rational allocation of medical resources in short supply during the process of dealing with the COVID-19 pandemic.

Although the function of each non-structural/accessory proteins of the SARS-CoV-2 is not yet fully understood, their protein sequences are highly similar to those of SARS-CoV. Most non-structural proteins always locate in the core of virion and play important roles in the pathogenesis. For example, RdRp, also called NSP12 of SARS-CoV, can catalyze the synthesis of viral RNA and plays an important role in the replication and transcription cycle of the virus ^28,29^. RdRp itself performs the polymerase reaction with limited efficiency, whereas NSP7 and NSP8 as co-factors can significantly stimulate its polymerase activity ^28^. Previous studies based on cryogenic electron microscopy (cryo-EM) indicated that the viral polymerase (RdRp-NSP7-NSP8 complex) might be excellent targets for developing new therapeutics of SARS and COVID-19 ^29,30^. NSP1 of the SARS-CoV may promote viral gene expression and immune escape by affecting interferon-mediated signal transduction ^31^. NSP4 is a multichannel membrane protein, which is an essential protein for viral replication ^32^. NSP9 plays a role of dimeric ssRNA binding protein during viral replication ^33,34^. NSP10 interacts with NSP14 and regulates ribose-2’-O-MTase activities involved in mRNA capping ^34-36^. In this study, we also found that IgG antibodies to non-structural/accessory proteins were positively correlated with routine clinical biomarkers of disease severity (procalcitonin, C-reactive protein, lactate dehydrogenase, D-dimer, IL-2R, and IL-6), but negatively correlated with the lymphocyte count. Therefore, nonsurvivors might result in more deaths of virus-infected cells and larger release of viral components from the dying cells than survivors, especially within 20 days after the symptom onset. Consequently, fuller interaction between viral non-structural/accessory proteins and the immune system of nonsurvivors resulted in stronger IgG responses to these proteins as evidenced in this study, which might underline the scientific background of these IgG responses as predicting signatures for the clinical outcome.

Moreover, some studies reported that treatment of COVID-19 patients with convalescent plasma was effective ^37,38^, whereas others did not observe the positive results ^39,40^. Several patients developed chills, rashes, shortness of breath, cyanosis, and severe dyspnea after treatment with convalescent plasma ^41^, which might attribute to the ADE. There are two distinct mechanisms of ADE occurrence during viral infections: (1) enhanced antibody-mediated virus uptake into Fcγ receptor IIa-expressing phagocytic cells thus leading to increased viral infection and replication; (2) excessive antibody Fc-mediated effector functions or immune complex formation causing enhanced inflammation and immunopathology ^11^. IgG antibodies against these non-structural proteins and accessory proteins might play important roles in the ADE during SAS-CoV-2 infections. To mitigate the potential risks of ADE with convalescent plasma therapy, plasma donors should be purified from donated convalescent plasma to enrich for neutralizing antibodies or monitor the levels of these IgG antibodies and to avoid the risks of ADE caused by non-neutralizing antibodies against these non-structural proteins. Currently, most of the COVID-19 subunit vaccines, such as mRNA-1273 and BNT162b2, are designed based on the S protein. In the present study, S1 specific IgG response is not a suitable predictor of the risk of COVID-19 mortality, which indicates the safety of these vaccines and is supported by the results of phase III clinical trials ^6,7^.

In conclusion, we provided a novel application of SARS-CoV-2 proteome microarray to detect serum IgG responses for early predicting COVID-19 death. Our results demonstrate that high level of IgG responses against 8 non-structural proteins and 2 accessory proteins on admission increased the COVID-19 mortality risk. Our research might improve clinical management and guide the development of effective medical interventions and vaccines by deeply understanding of the pathogenesis of COVID-19.

## Data Availability

The data used to support the findings of this study are available from the corresponding author upon request.

## Acknowledgments

We thank Prof. H. Eric Xu (Shanghai Institute of Materia Medica) for providing RdRp protein. We also thank Healthcode Co., Ltd., Hangzhou Bioeast biotech Co., Ltd. and Vacure Biotechnology Co.,Ltd. for providing the proteins.

## Author Contributors

X-L.F., S-C.T., and F.W. performed experiments and designed the study. Q.L., C-Z.Y., and Y.L. performed the experiments. H-Y.H., Z-Y.S., and B.Z. collect specimens. D-Y.L., J-B.X., Z-Q.O., J-L.B., and Z-W.X. prepared the reagents. Y-D.Z., Z-J.Y., H.C., Y-X.Z., and X-S.L. analyzed the data. M-A.H, X-N.W., H-W.J., H-N.Z., H.Q., and S-J.G. took responsibility for the accuracy of the data analysis. X-L.F., Q.L., and C-Z.Y. wrote the manuscript with suggestions from other authors.

